# A Hybrid Machine Learning Framework to Predict Early Risk of Mortality in Paralytic Ileus Patients using Electronic Health Records

**DOI:** 10.1101/19006254

**Authors:** Fahad Shabbir Ahmad, Liaqat Ali, Raza-Ul-Mustafa, Hasan Ali Khattak, Syed Ahmad Chan Bukhari

## Abstract

**Background and Objective:** Paralytic Ileus (PI) is the pseudo-obstruction of the intestine secondary to intestinal muscle paralysis. PI is caused by several reasons such as overuse of medications, spinal injuries, inflammation, abdominal surgery, etc. We have developed an early mortality prediction framework that can help intensivist, surgeons and other medical professionals to optimize clinical management for PI patients in terms of optimal treatment strategy and resource planning.

**Methods:** We used publicly available ICU database called MIMIC III v1.4, extracted patients that had paralytic ileus as primary diagnosis over the age of 18 years old. We developed FLAIM Framework a two-phase model (Phase I: Statistical testing and Phase II: Machine Learning application) that was compare to traditional methods of machine learning. We used five different machine learning algorithms to test the validity of our Framework. We evaluated the effectiveness of the proposed framework by comparing accuracy, sensitivity, specificity, Receiver Operating Characteristic (ROC) curves, and area under the curve (AUC) for each model.

**Results:** The highest improvement in AUC of 7.78% was observed due to application of the proposed FLAIM method. Additionally, almost for all the machine learning models, improvement in accuracy was also observed. With the FLAIM framework, we recorded an accuracy of 81.30% and AUC of 81.38% under support vector machine (with RBF kernel) model in predicting mortality during a hospital stay for the PI patients

**Discussion:** Our results show promising clinical outcome prediction and application for individual patients admitted to the ICU with paralytic ileus after the first 24 hours of admission.

## 1. Introduction and Background

Paralytic ileus (PI) or intestinal pseudo-obstruction is a rare disorder of the intestine which is characterized by the paralysis of the intestinal peristalsis without any machinal cause. This leads to an impaired movement of food or fecal matter along the gastrointestinal tract [1]. Diagnosis is made clinically and it can present with symptoms like abdominal bloating, pain, nausea, vomiting and constipation as initially described by Dudley et al [2]. PI is caused by either primary or idiopathic dysfunction of gastrointestinal motility (acquired or inherited mutations) [3]. Additionally, it could be due to abdominal and/or pelvic surgery [4], systemic lupus erythematosus [5], scleroderma [6], Parkinson’s disease [7], infections [8], medications like opioids and antidepressants [9], radiation to the abdomen [10] and malignancies lung cancer [11]. Unfortunately, paralytic ileus has a poor clinical outcome [12] and an early prediction could be helpful for care delivery.

The mortality of patients with PI can be as high as 40% [13] in the ICU setting. Patients admitted to the intensive care are especially at risk of dying because of the seriousness of their condition. Some of these ICU may have additional risks associated when these patients undergo colonic decompression [14] and while providing bowel rest with reduced food intake [15]. With this in mind, it becomes important to have an approach that can predict the risk of mortality in these patients and help clinicians in making a decision faster and allocate adequate resources to these patients.

The MIMIC-III (Medical Information Mart for Intensive Care III) database is a freely-available. large database that has de-identified patient data associated with more than forty thousand patients that were admitted to the critical care units of the Beth Israel Deaconess Medical Center between 2001 and 2012 and developed by the Massachusetts Institute of Technology (MIT) [16]. This dataset has been used to predict mortality for multiple diseases [17]. In this paper, we utilized risk factors that are based on laboratory test results in order to predict mortality in PI patients.

To obtain an optimal mortality risk prediction performance, we proposed an automated framework called FLAIM. The proposed FLAIM framework works in two phases. In the first phase, it uses Cox-regression (univariate and multivariate) and Kaplan-Meier analysis based statistical methods to find out a relevant (reduced) set of risk factors while the second phase explores the feasibility of different machine learning models which are used for classification purposes. Machine learning models are provided with some training data, the model learns a hypothesis i.e. a fitting function denoted by h(x) by optimizing error achieved on the training data. Thus, the model learns the behavior of the distribution of the data by analyzing the training data set. The model performance i.e. the performance of the learned hypothesis function is tested on the unseen testing data. The main objective is to construct such a model that would show better performance on the training data and would also generalize to unseen data i.e. the testing data. Hence, such a constructed model can be deployed in hospitals for predictions in future.

In this paper, we have explored the hybridization of six different machine learning models with the above-mentioned statistical models that are utilized for evaluating most relevant (reduced) set of risk factors. The machine learning models used for mortality prediction of PI patients include linear discriminant analysis (LDA), (GNB), Decision Tree (DT) model (also known as CART in literature), K nearest neighbors (KNN) and support vector machines (SVMs) with linear and radial basis functions (RBF). Linear Discriminant Analysis (LDA) is a method used in statistics, pattern recognition and machine learning (ML) to find a linear combination of features which characterizes or separates two or more classes of objects or events [18]. SVM has been used in bioinformatics fields for its excellent performance in classification by drawing a hyperplane [22] [23]. A hyperplane is a line that divides two classes where each labeled data lay on one side. To separate classes, there may be many possible hyperplanes that can differentiate instances, however, the goal is to find an optimal hyperplane that has a maximum margin or maximum distance from the data points of both classes [20,24]. When the samples are not separable then an error term is considered as cost function that leads to poor classification. Regularization methods are used to deal poor classification issues such as overfitting and underfitting.

In addition to the classification models stated above, in FLAIM we have also used Decision Tree model. A decision tree has nodes (attributes) and edges which make a machine processable tree-like structure which helps to build a parseable logic that leads to a decision, where edges are the outcomes of the split to the next node. The root node performs the first split, whereas leaves, terminal nodes predict the outcome. Entropy and information gain are the mathematical methods that have been used for the splitting of nodes [25]. Another machine learning model used for mortality prediction of PI patients is GNB model. It uses Bayes theorem of probability for prediction of unknown class. Usually, three approaches have been used for classification in Naive Bayes. 1) Gaussian: It assumes that continuous features follow a normal distribution, 2) Multinomial: When features are discrete, 3) Bernoulli: When features are binary [26,27][28] is most frequently non-parametric methods used in data science. In kNN, (*k*) is the number of nearest neighbors where unknown instances or data-points are classified based on the nearest known instances in the feature space. The instances which are close to each other are more likely of the same class. Distance between instances is commonly measured using the Euclidean distance [29]. The principal of kNN during training is to store all data points then the prediction algorithm calculates the distance from new data-point (*x*) to all points in the data and finally assigns the instance to that class which has the shortest distance.

## 2. Methods

### 2.1 Study Type

Retrospective investigation of clinical electronic health records

### 2.2 Participants

The data was obtained from a publicly available database to researchers called MIMIC III. The was collected by many different intensive care units including surgical, trauma surgery, medical, coronary and cardiac surgery at Beth Israel Deaconess Medical Center (BIDMC), Boston, Massachusetts from 2001-2012. All patients’ records are completely anonymous, and all the data collected has received a prior Institutional Review Board (IRB) approval from BIDMC and Massachusetts Institute of Technology (MIT). The institutional IRB requirement was waived because the database is HIPAA-complaint deidentified and our study complies with the ethical standards set by the Helsinki declaration (1964) and its amendments [16]. This database contains clinical parameters from de-identified ICU admission patients to the above mentioned.

We selected all patients using the ICD-9 code for paralytic ileus (560.1) [30] from all and extracted all the data from the rest of the datasets including initial labs, demographic information, ICU stay data, microbiological reports, and prescription information. We excluded calculated the age using the date of birth and date of admission and remove all patients that were under the age of 18 for this analysis.

### 2.3 Testing methods

Initial labs for the first day of ICU admission were extracted from the data set using the MIMIC code available for public access repository on GitHub, which included hemoglobin, hematocrit, white blood cells, platelets, serum electrolytes (sodium, potassium, chloride, bicarbonate), blood urea nitrogen (BUN), creatinine, glucose, anion gap, lactate, bilirubin, albumin, prothrombin time (PT), partial-thromboplastin time (PTT) and international normalization ratio (INR). All of these labs were expressed as minimum and maximum values for each blood marker, we calculated the average and subdivided the values into either normal range and abnormal values. Reference ranges were used to calculate these values and can be seen in Table 2.

### 2.4 Analysis

We have developed the FLAIM (Fahad-Liaqat-Ahmed Intensive Model) a machine learning framework for the early mortality risk prediction of patients with the PI disease. Our proposed framework works in two phases (Figure 1).

**Figure 1.**
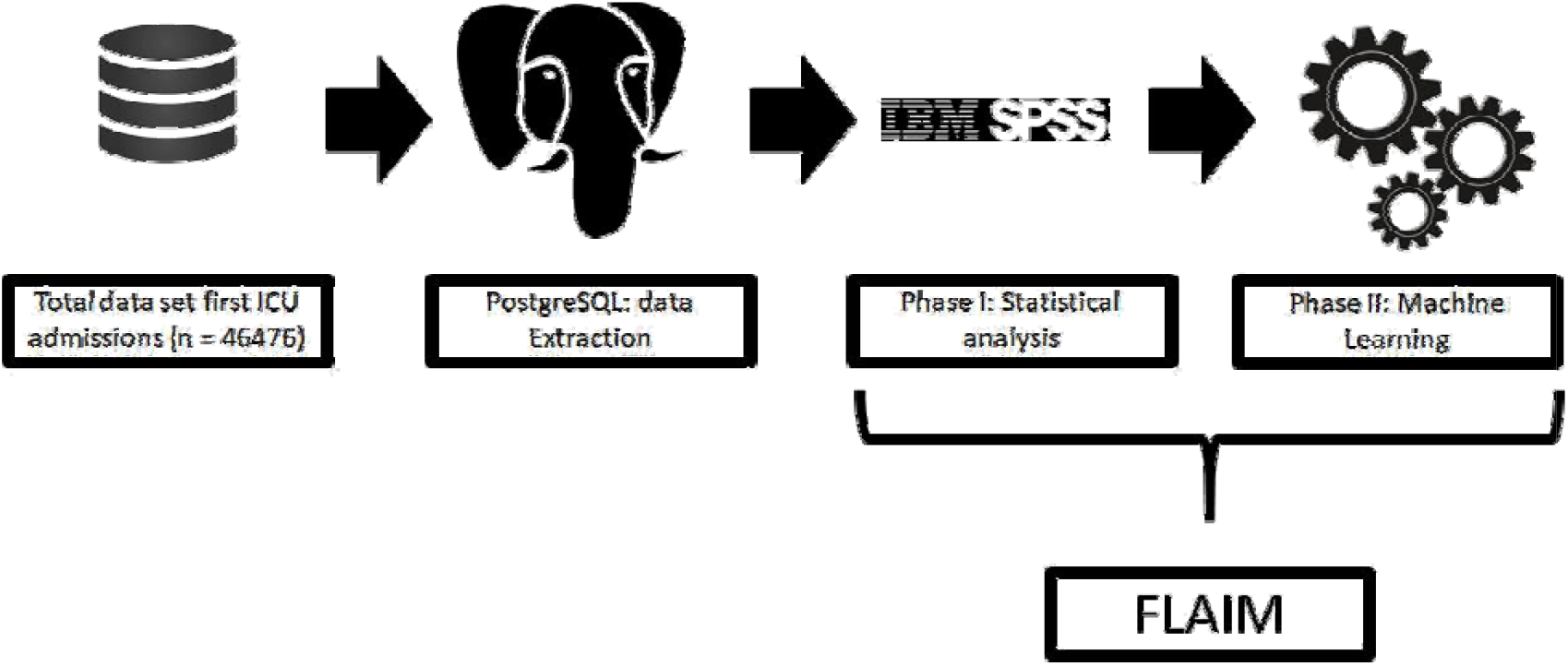
Consort diagram and the FLAIM Framework elaboration.

This proposed hybrid model can be considered as a black-box that hybridizes the risk survival statistical methods with machine learning-based predictive models. The first stage/phase is significant as it improves the predictive capabilities of machine learning-based predictive models (phase II) as well as reduces the time complexity of the predictive models by reducing the number of risk factors that need to be processed by the machine learning models.

#### Phase I

For the initial data extraction, we used PostgreSQL (version 11.2, www.postgresql.org) and data analysis was done with IBM SPSS (version 24.0.0.0) [31] for demographic, univariate, multivariate and Kaplan Meier curves for outcome assessment. A p-value of <0.05 was considered statistically. Statistical analysis is carried out on all the variables, this includes univariate and multivariate (covariates with age, gender, and ethnicity) cox-regression analysis followed by Kaplan-Meier survival analysis (Figure 2).

**Figure 2.**
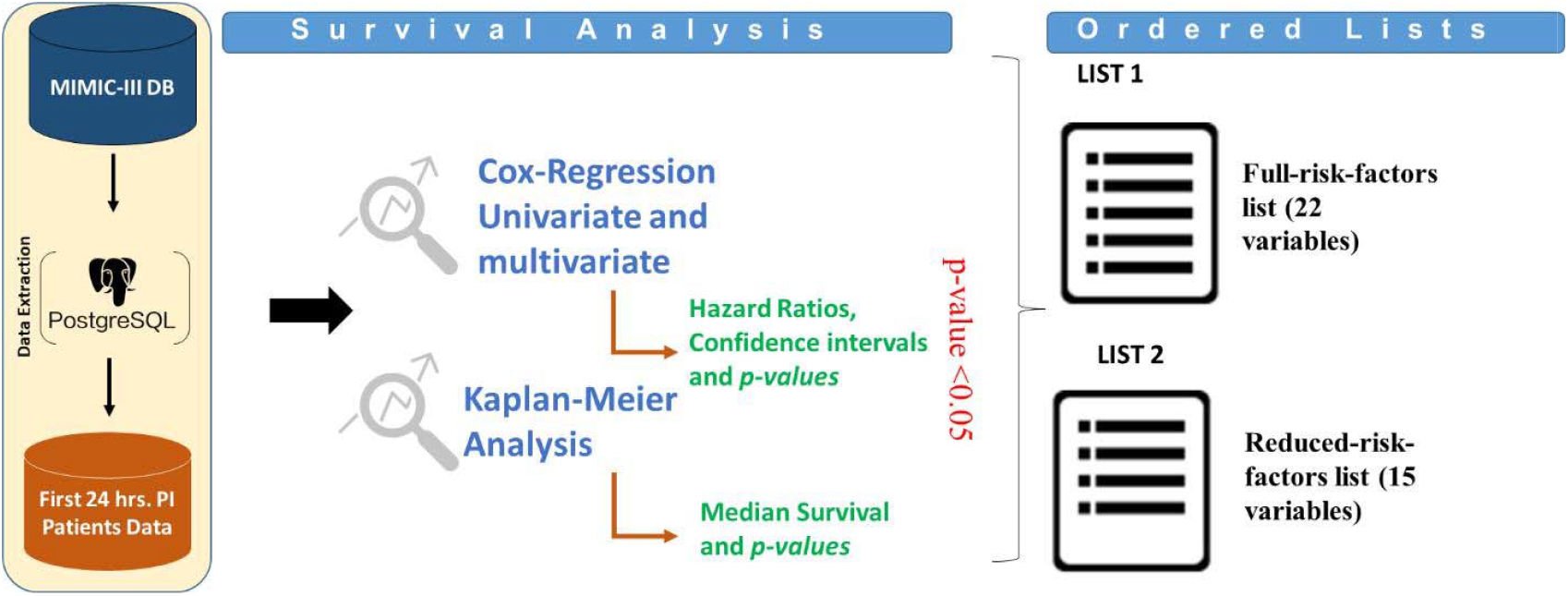
FLAIM Phase I (statistics phase). once day is extracted from MIMIC-III database, univariate, multivate Cox-regression analysis is done followed by a Kaplan Meier analysis to generate two lists of risk factors i.e. full-risk-factors list (22 variables) and reduced-risk-factors list (15 variables) based on the statistics that were performed.

The cox-regression analysis [32] gives us the hazard ratio for these risk factors which can be used in conjunction with the Kaplan-Meier analysis [33] to make a rank order list of these variables ranked from highest to lowest; two lists were generated full-risk-factors list (22 variables) and reduced-risk-factors list (only statistically significant results were used, 15 variables). The reduced set of risk factors that are obtained based on the statistical methods, they are supplied to different machine learning models for classification. The outcome of this phase can be viewed in Figure 3.

**Figure 3.**
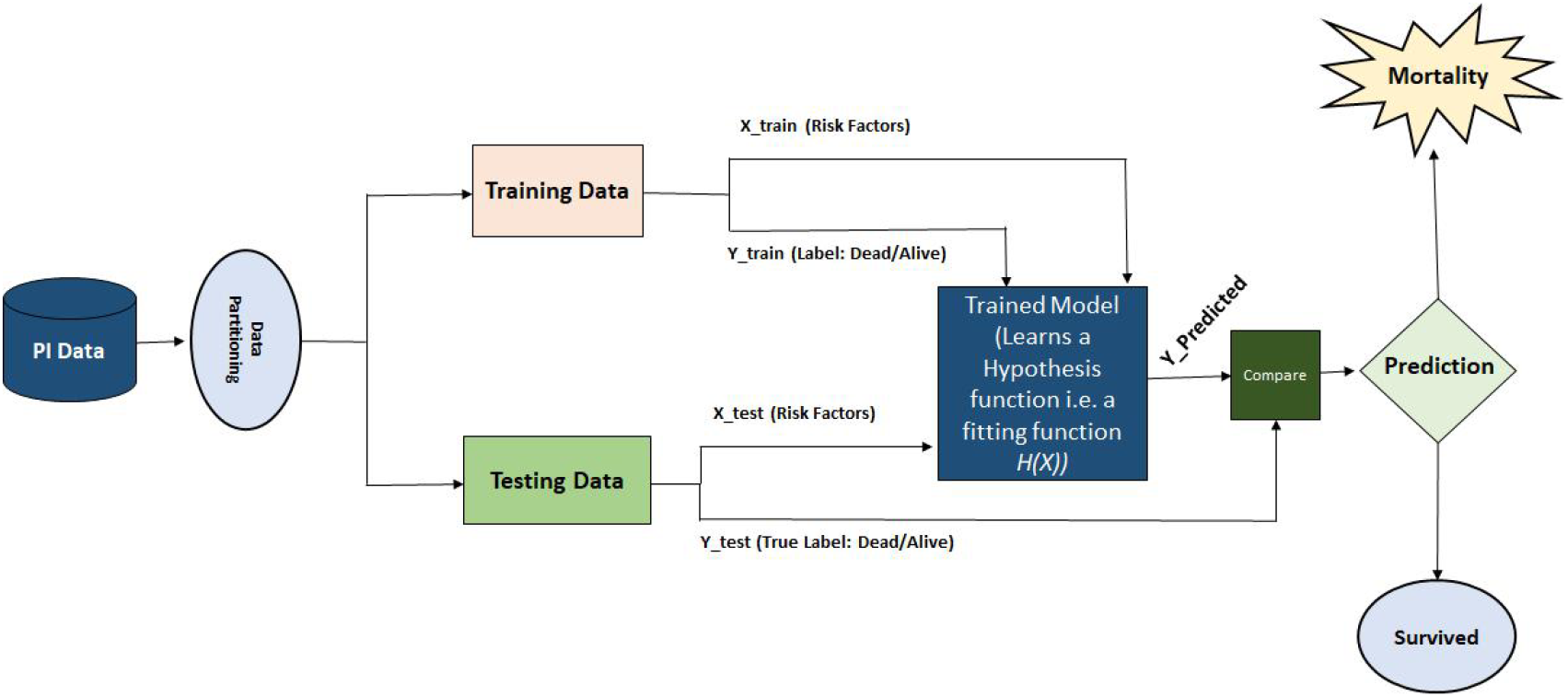
FLAIM Phase II machine learning.

#### Phase II

In this phase, different machine learning models are applied to risk factors that have been found significant in Phase I, and higher weight can be given to the variables/risk factors based on hazard ratios as shown in Table 3. During the second phase, the data (risk factors) is partitioned in two parts i.e., training and testing datasets (Figure 4). The training dataset with risk factors or feature vectors denoted by X_train along with the label information denoted by Y_train are supplied to the machine learning models. The machine learning models analyze the distribution of the training data and tries to learn a hypothesis function i.e., a fitting function that would represent the behavior or distribution of the whole dataset. In the next step, the performance of the trained or learned machine learning model is tested by considering the testing dataset. During the testing process, only feature vector or risk factors denoted by X_test is supplied to the trained or learned machine learning model, the model predicts the label based on the learned knowledge or rules from the training data. The predicted labels are compared with the true labels of samples of the testing dataset and based on the comparison, the prediction accuracy is evaluated.

**Figure 4:**
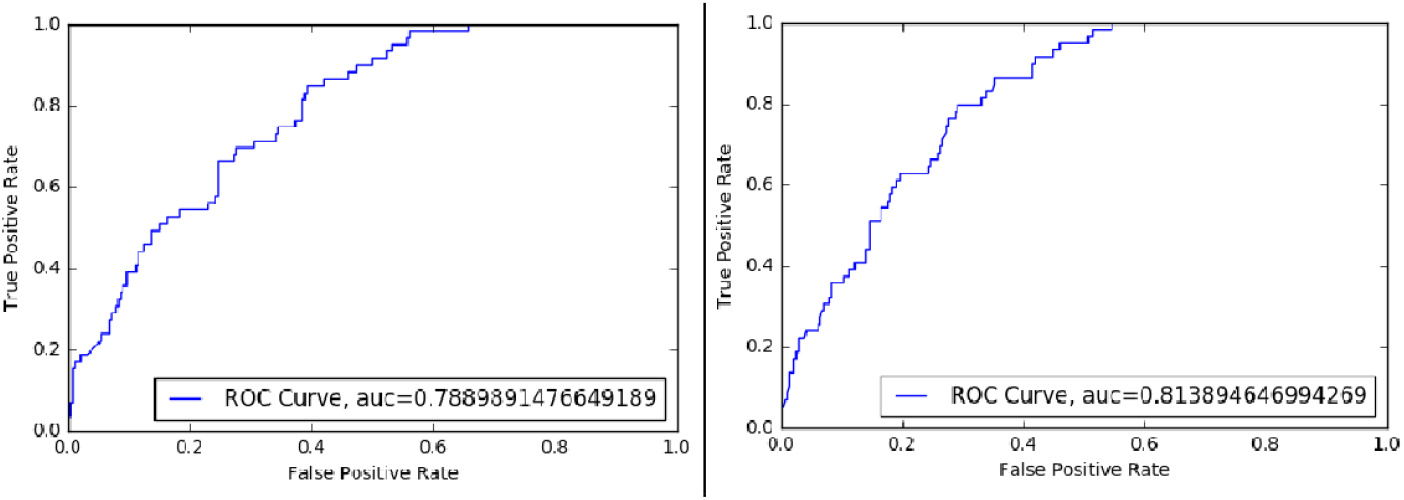
Left ROC curve is of conventional SVM RBF and right ROC curve is that of SVM RBF model developed under FLAIM architecture.

## 3. Results

### 3.1 Participants

We used the complete list of risk factors (full list) and a selected list of risk factors (reduced list) (Table 3.) to predict outcome in the test-set using Linear Discriminant Analysis, Gaussian Naive Bayes, Decision Trees, K-Nearest neighbor and support vector machines (Supp 1.) In order to evaluate the effectiveness of the proposed method, we used the publicly available MIMIC III dataset. A total of 1021 patients’ data were extracted from the original dataset of the MIMIC III database that were diagnosed with PI. All the patients were above the age of 18 years of age at their first admission. The median age of patients was 63.2 years, most of the people that were admitted to the ICU were women (63.8%) and mostly Caucasian (74%) from all the ethnic groups. These patients had a median stay of 14.5 ± 16.2 days of hospital stay and 4.2 ± 10.7 days of intensive care stay. Emergency admission comprised of most admission making 76.7% of all patients that were admitted to the ICU while most admissions came through referrals around 490 patients (48%). Mortality among these patients was 16.8% (173), while 441 (43.2%) were transferred out to different health facilities and 407 (40%) patients were discharged home. All of this can be seen in Table 1.

**Table 1.** Patient demographics

| Demographics |  |  |  |
| --- | --- | --- | --- |
| | | Median ± SD | Range |
| Age | | 63.2 ± 15.4 years | 19-89 years |
| Hospital stay (days) | | 14.5 ± 16.2 days | 0-133 days |
| ICU stay (days) | | 4.2 ± 10.7 days | 0.03-57.4 days |
|  |  | n | % |
| Gender | Male | 370 | 36.2 |
|  | Female | 651 | 63.8 |
| ICU type on admission | CCU | 60 | 5.9 |
|  | CSRU | 125 | 12.2 |
|  | MICU | 452 | 44.3 |
|  | SICU | 215 | 21.1 |
|  | TSICU | 153 | 15 |
| Admission Type | Elective | 238 | 23.3 |
|  | Emergency | 783 | 76.7 |
| Admission Location | Referrals | 490 | 48 |
|  | Transfers | 197 | 19.3 |
|  | ER admissions | 334 | 32.7 |
| Discharge to | Health care facility | 441 | 43.2 |
|  | Home | 407 | 40 |
|  | Dead | 173 | 16.8 |
| Ethnicity | African American | 93 | 9.1 |
|  | Asian | 28 | 2.7 |
|  | Hispanic | 31 | 3 |
|  | Others | 29 | 2.8 |
|  | Unknown | 84 | 8.2 |
|  | Caucasian | 756 | 74 |

**Table 2.** All the variables that were used in these analyses and the reference ranges we used as cut-off points to distinguish between normal and abnormal levels. Also show the univariate and multivariate analysis of these serum markers with their confidence intervals. * = significant (*p*-value <0.05). WBC = white blood cells, PTT = Partial Thromboplastin Time, PT = Prothrombin and INR = International Normalization Ratio.

| Labs |  | Reference ranges | Units | n (abnormal levels) | % | Univariate |  |  | Multivariate |  |  |
| --- | --- | --- | --- | --- | --- | --- | --- | --- | --- | --- | --- |
|  |  |  |  |  |  | *p*-value | HR | 95%CI | *p*-value | HR | 95%CI |
| Chemistry | Sodium | 135-145 | mEq/L | 269 | 26.3 | 0.041* | 1.39 | 1.02-1.89 | 0.008* | 1.39 | 1.02-1.89 |
|  | Potassium | 3.5-5.5 | mmol/L | 120 | 11.8 | 0.025* | 1.65 | 1.09-2.49 | 0.005* | 1.67 | 1.11-2.53 |
|  | Chloride | 97-107 | mEq/L | 430 | 42.1 | 0.993 | 1.00 | 0.74-1.35 | - | - | - |
|  | Bicarbonate | 23-29 | mEq/L | 524 | 51.3 | 0.001* | 1.69 | 1.22-2.33 | <0.001* | 1.73 | 1.25-2.39 |
|  | Blood urea nitrogen | 7-20 | mg/dL | 543 | 53.2 | <0.001* | 3.67 | 2.45-5.51 | <0.001* | 2.38 | 2.38-5.39 |
|  | Creatinine | 0.6-1.2 | mg/dL | 490 | 48 | <0.001* | 2.11 | 1.53-2.93 | <0.001* | 2.34 | 1.61-3.11 |
|  | Glucose | 72-99 | mg/dL | 895 | 87.7 | 0.014* | 0.59 | 0.39-0.88 | 0.002* | 0.53 | 0.35-0.80 |
|  | Anion Gap | 8-16 | mEq/L | 246 | 24.1 | <0.001* | 2.71 | 2.01-3.65 | <0.001* | 2.77 | 2.04-3.76 |
|  | Lactate | 0.5-1.0 | mmol/L | 675 | 66.1 | 0.003* | 2.72 | 1.30-5.82 | 0.008* | 2.63 | 1.22-5.65 |
|  | Bilirubin | 0.1-1.2 | mg/dL | 234 | 59 | 0.000* | 1.81 | 1.31-2.53 | <0.001* | 1.93 | 1.38-2.71 |
| Hematological | Hematocrit | 37-52 | % | 847 | 83 | 0.696 | 0.92 | 0.60-1.41 | - | - | - |
|  | Hemoglobin | 13.5-15.5 | g/dL | 940 | 92 | 0.931 | 1.03 | 0.51-2.10 | - | - | - |
|  | WBC | 4.5-11 | 1000 cells/mL | 585 | 57.3 | 0.066 | 1.34 | 0.98-1.85 | - | - | - |
|  | Platelets | 150-350 | 1000 cells/mL | 417 | 40.8 | 0.007* | 1.52 | 1.12-2.07 | 0.001* | 1.64 | 1.20-2.23 |
| Bleeding profile | PTT | 25-35 | Seconds | 467 | 45.7 | 0.002* | 1.65 | 1.20-2.28 | 0.003* | 2.63 | 1.20-2.28 |
|  | PT | 11-13.5 | Seconds | 645 | 63.2 | <0.001* | 2.55 | 1.62-4.04 | <0.001* | 1.66 | 1.67-4.16 |
|  | INR | 0.8-1.1 |  | 726 | 71.1 | 0.020* | 1.79 | 1.1-2.92 | 0.009* | 1.81 | 1.11-2.95 |
| Albumin |  | 3.5-5.5 | g/dL | 398 | 39 | 0.386 | 0.80 | 0.50-1.30 | - | - | - |

**Table 3.** Performance of different machine learning models on the current dataset. LDA = linear discriminant analysis, GNB = gaussian naive bayes, KNN = k-nearest neighbor, SVM = support vector machine, Lin = linear and RBF = radial basis functions.

| Method | AUC | Training Accuracy (%) | Testing Accuracy (%) | Sensitivity | Specificity |
| --- | --- | --- | --- | --- | --- |
| **LDA (Baseline)** | 0.774 | 85.53 | 84.57 | 83 | 58 |
| **LDA (Proposed)** | 0.817 | 84.94 | 84.87 | 90 | 63 |
| **GNB (Baseline)** | 0.737 | 77.92 | 76.86 | 90 | 63 |
| **GNB (Proposed)** | 0.798 | 76.75 | 78.64 | 84 | 62 |
| **Decision Tree (Baseline)** | 0.554 | 100 | 74.48 | 24 | 87 |
| **Decision Tree (Proposed)** | 0.626 | 98.09 | 75.67 | 40 | 86 |
| **KNN (Baseline)** | 0.752 | 85.97 | 83.09 | 78 | 34 |
| **KNN (Proposed)** | 0.790 | 85.97 | 85.46 | 83 | 65 |
| **SVM (Lin) (Baseline)** | 0.636 | 83.33 | 82.49 | 59 | 60 |
| **SVM (Lin) (Proposed)** | 0.810 | 84.65 | 83.38 | 90 | 63 |
| **SVM (RBF) Model (Baseline)** | 0.673 | 83.33 | 82.49 | 80 | 38 |
| **SVM (RBF) Model (Proposed)** | 0.810 | 84.65 | 83.38 | 90 | 63 |

The univariate analysis showed that patients with abnormal blood levels for sodium, potassium, bicarbonate, BUN, creatinine, glucose, anion gap, lactate, bilirubin, platelets, PT, PTT, and INR had worse survival than those that had normal ranges of these blood related risk factors or biomarkers. The multivariate analysis was also done so that we normalize these findings for age, gender and ethnicity and all these markers were still significant (Table 2.)

### 3.2 Analysis Participants

Kaplan-Meier survival analysis of these blood markers revealed that at the first 24 hours of ICU admission labs show a relationship between better overall survival during these patient’s hospital stay. The largest difference between median patient survival was seen with abnormal vs normal anion gap (37 vs 129 days(d)) a difference of 92 days. Serum abnormal vs normal bicarbonate (55d vs 129d), PT (55 vs median not reached (MNR)), BUN (51d vs MNR), bilirubin (37d vs 67d), creatinine (57d vs 78d), PTT (54d vs 67d) and platelets (55d vs 66d) were also significant (Table 2, Figure 4 and Figure 5). Showing abnormalities of these markers lead to worse patient survival during their hospital stay in these patients.

These results were used to make an ordered list of risk factors that affect patient outcomes from most to least as described in table 3. We used previously established machine learning algorithms as shown in supplemental 1.

Subsequently, we used two third (2/3) i.e., n=684 samples for training the machine learning models while one third of the dataset (n=337) for testing purposes. The results for each machine learning model is presented in Table 3. Our assessment of the algorithm was based on testing set accuracy sensitivity, specificity and area under the ROC curve (AUC) to assess for the best algorithm. As in machine learning, the quality of output of a model is judged by AUC, hence, we utilize the AUC and accuracy as the primary evaluation metrics for selecting optimal model under the proposed FLAIM architecture. Our best algorithm SVM RBF on reduced risk factors yielded highest AUC of 81.38%, testing accuracy of 81.30%, training accuracy of 83.18%, sensitivity of 35.59%, a specificity of 91%. The second-best algorithm was LDA with AUC of 82.02%, testing accuracy of 70.32%, training accuracy of 69.59%, sensitivity of 83%, a specificity of 58% using the reduced set of risk factors.

## 5. Discussion

Paralytic ileus although a rare but a morbid and fatal condition [34] and almost 16.8% mortality to all patients that were admitted to the ICU in our patient population. For that reason, it becomes important to identify patients that may be at risk of dying during their ICU stay. Out dataset revealed that the majority of admitted patients are elderly individuals with a median age of 63 years [34–36]. In this cohort of patients, we observed labs which were performed in the first 24 hours to predict overall survival. The best predictors of patient survival were serum anion gap (37d vs 129d), bicarbonate (55d vs 129d), PT (55d vs MNR), BUN (51d vs MNR), bilirubin (37d vs 67d), creatinine (57d vs 78d), PTT (54d vs 67d) and platelets (55d vs 66d). While other labs like serum chloride, hematocrit, hemoglobin, white blood cells, and albumin were non-significant. The abnormal initial anion gap is associated with adverse outcomes in admitted to the ICU [37] which is also seen in the study as well. Similarly, abnormal potassium[38] bicarbonate [39], BUN [40], bilirubin [41], creatinine [42], platelets [43]. Abnormal coagulation studies in a patient with sepsis have shown worse outcomes in ICU patients. We considered patients with age > years along with their demographic information such as gender and ethnicity and prepared a final list of all risk factors and further ranked them according to their severity (Table 3).

In order to validate the effectiveness of the proposed hybrid model, we developed two types of experiments. In the first experiment, we utilized the conventional machine learning models such as LDA, GNB, DT, KNN, and SVM as baseline using full set of risk factors. In the second experiment, the same machine learning models were hybridized with the statistical models using the proposed FLAIM method (see Figure 1). The main objective of developing two different types of experimental settings was to validate the fact that the proposed hybrid method is capable of reducing the complexity of machine learning models as well as enhancing their predictive performance. The performance comparison or evaluation of the two experiments were carried out from two aspects i.e. using mortality prediction accuracy and ROC curve to yield AUC. Both the accuracy and AUC are well used metrics to evaluate a model performance. Therefore, these two-evaluation metrics were exploited for comparison of the proposed methods with baseline methods.

From Table 3, the improvement in AUC of conventional machine learning (baseline) models can be observed. The baseline models and their AUC have been reported in red and the AUC for the corresponding proposed or developed methods (under FLAIM architecture) have been reported in green color. It can be seen that the optimal performance is obtained with SVM RBF model as it yields accuracy rate of 81.30% and AUC of 81.38% which is better compared to the other baseline and proposed methods. Additionally, the constructed model has another benefit of lower complexity i.e., it uses reduced set of risk factors. This means if we provide risk factors (serum lactate, BUN, PT, anion gap, creatinine, INR, bilirubin, potassium, PTT, bicarbonate, platelets, sodium, glucose, Age more than 65 years and gender) the model will correctly predict ∼81% of the time if the patient will die during their hospital stay or not. Other models that showed potential similar results but were less accurate than the optimal model, were LDA (reduced risk factors), DT (reduced risk factors) and SVM (Linear) with full risk factors and reduced set of risk factors. To effectiveness of the proposed SVM (RBF) under FLAIM architecture is validated by observing the AUCs in Figure 4 of conventional SVM (RBF) and the proposed version of SVM (RBF). The AUC of conventional SVM RBF is 78.89% while the AUC of the proposed SVM RBF is 81.38%, hence, the improvement of 2.49% can be observed. Similarly, the ROC charts for each model (conventional and proposed) are depicted in Figure 3 (see supplementary section). For almost all the models, the improvement in AUCs can be observed due to application of the proposed FLAIM architecture. Hence, our proposed framework could potentially facilitate the critical care physicians in determining which patients are at the highest risk of mortality while their hospital stay and should be treated with more care.

Our future direction is to develop a more sophisticated model that incorporates more clinical parameters and has a higher AUC and accuracy of predicting clinical outcome for paralytic ileus patients admitted to the intensive care unit. Further experiments are required to evaluate the performance of the proposed framework with different age groups in different clinical situations.

## 6. Conclusion

FLAIM is a statistically robust machine learning framework which is developed for clinical use to predict mortality risk in PI patients. The mortality prediction was modelled based on clinically relevant labs or risk factors that were collected from the PI patients during their first 24-hours of stay in the intensive care unit. Experimental results evidently show that the proposed method reduces the complexity of conventional learning models by effectively reducing the number of risk factors and also improves the predictive performance and output quality of conventional machine learning models. From the simulation results, it can be safely concluded that the proposed method can also be expanded in the future to predict mortality and other risks associated with the ICU admitted patients for various diseases.

## Data Availability

MIMIC III database version 1.4

## Supplemental Figures and Table

**Supplemental Table 1.**
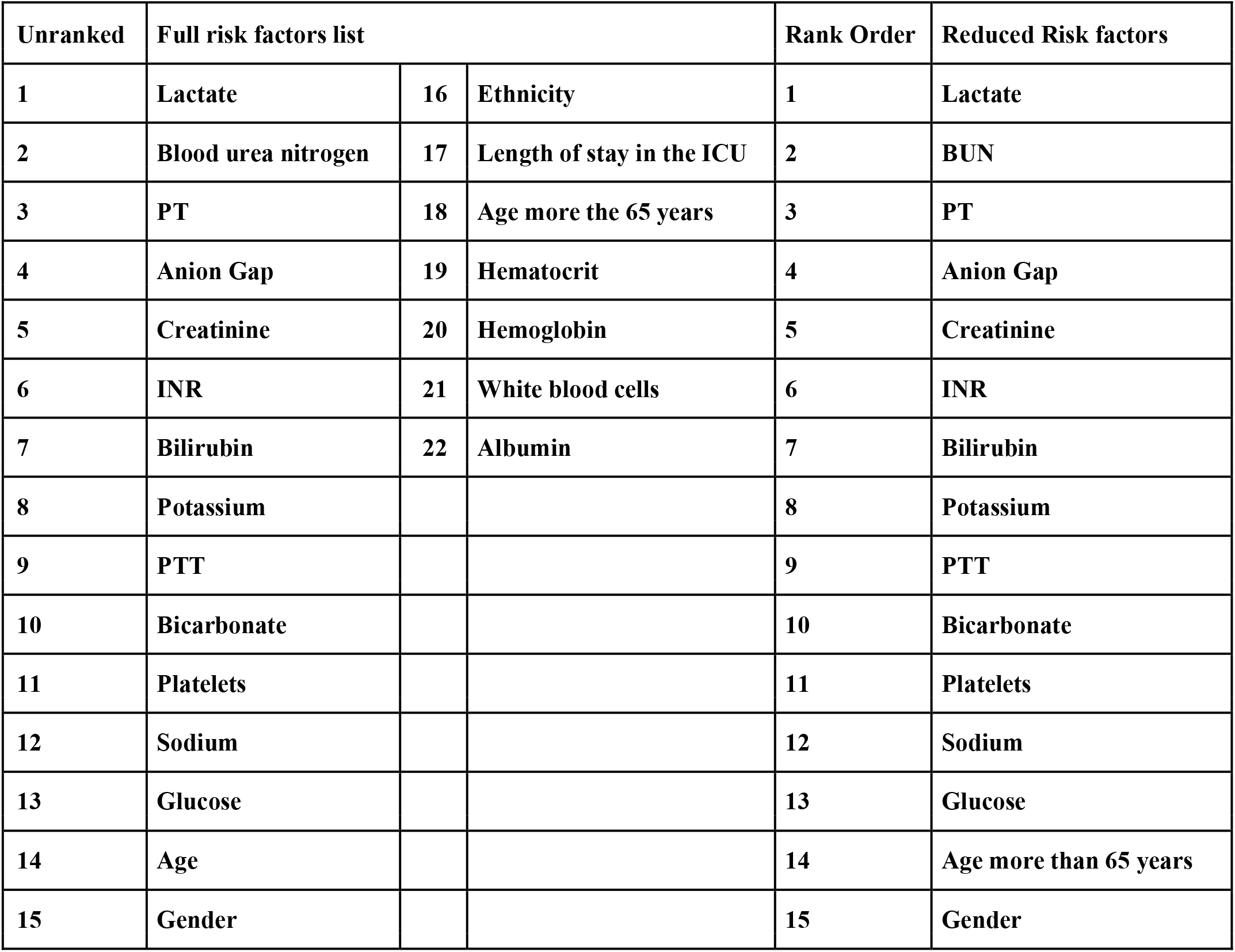
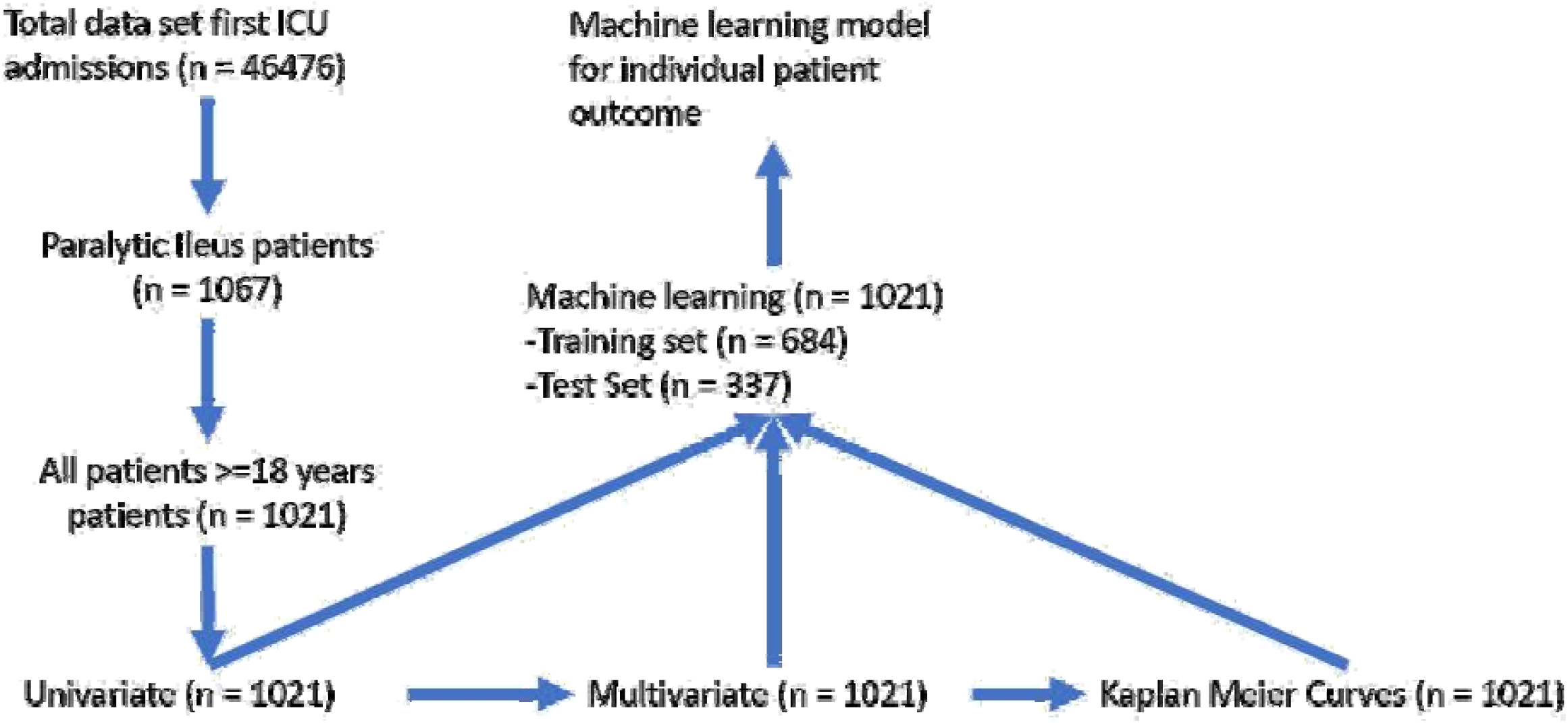
Risk factors list. 1 to 22 (full list of risk factors) and 1 to 15 (Reduced list of risk factors)

**Supplemental Figure 2.**
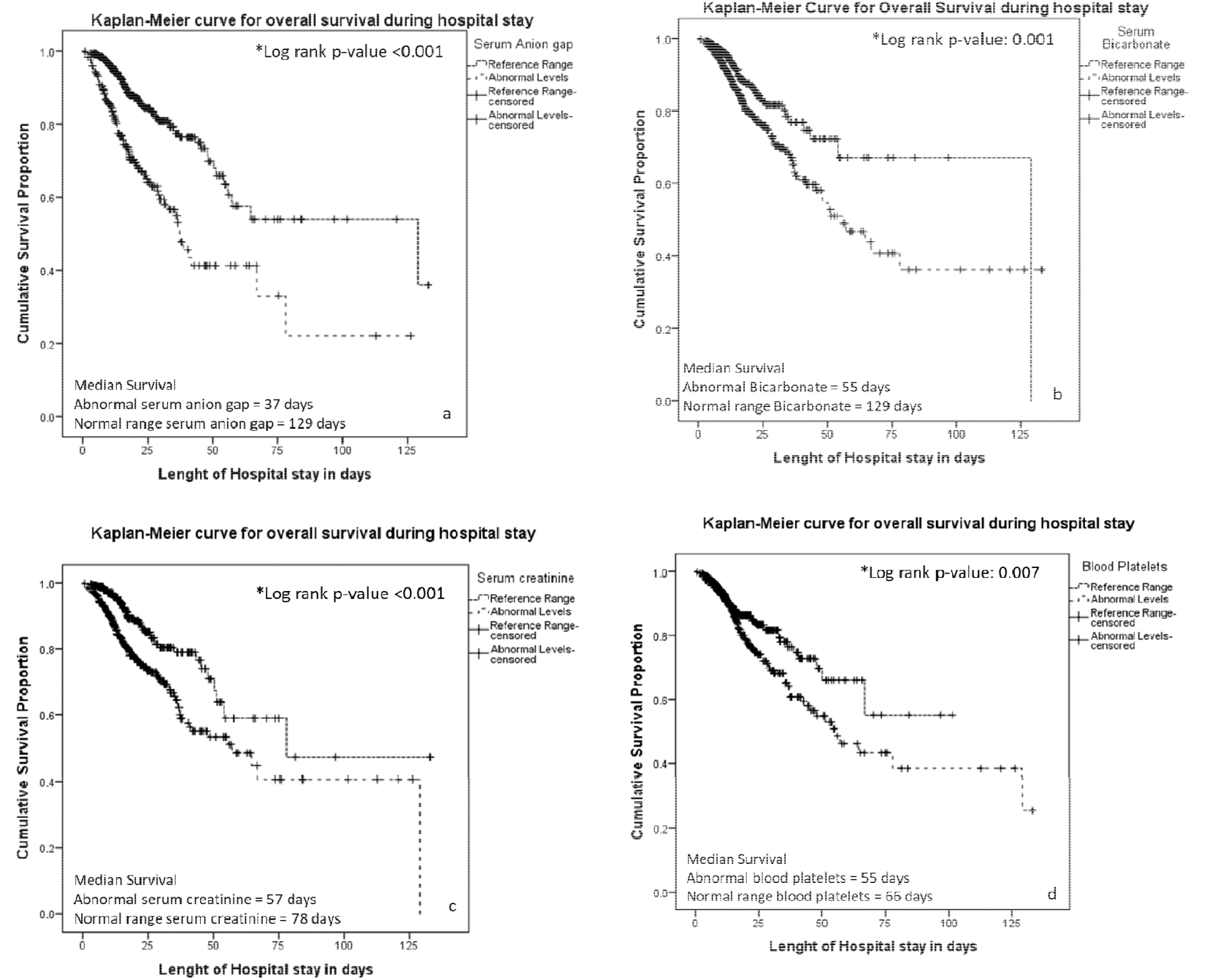
Kaplan-Meier Curves for a) abnormal serum anion gap (MS 37 days vs 129 day), b) abnormal serum bicarbonate (MS 55 days vs 129 day), c) abnormal serum (MS 57 days vs 78 day), d) abnormal serum (MS 55 days vs 66 day). MS = median survival

**Supplemental Figure 3.**
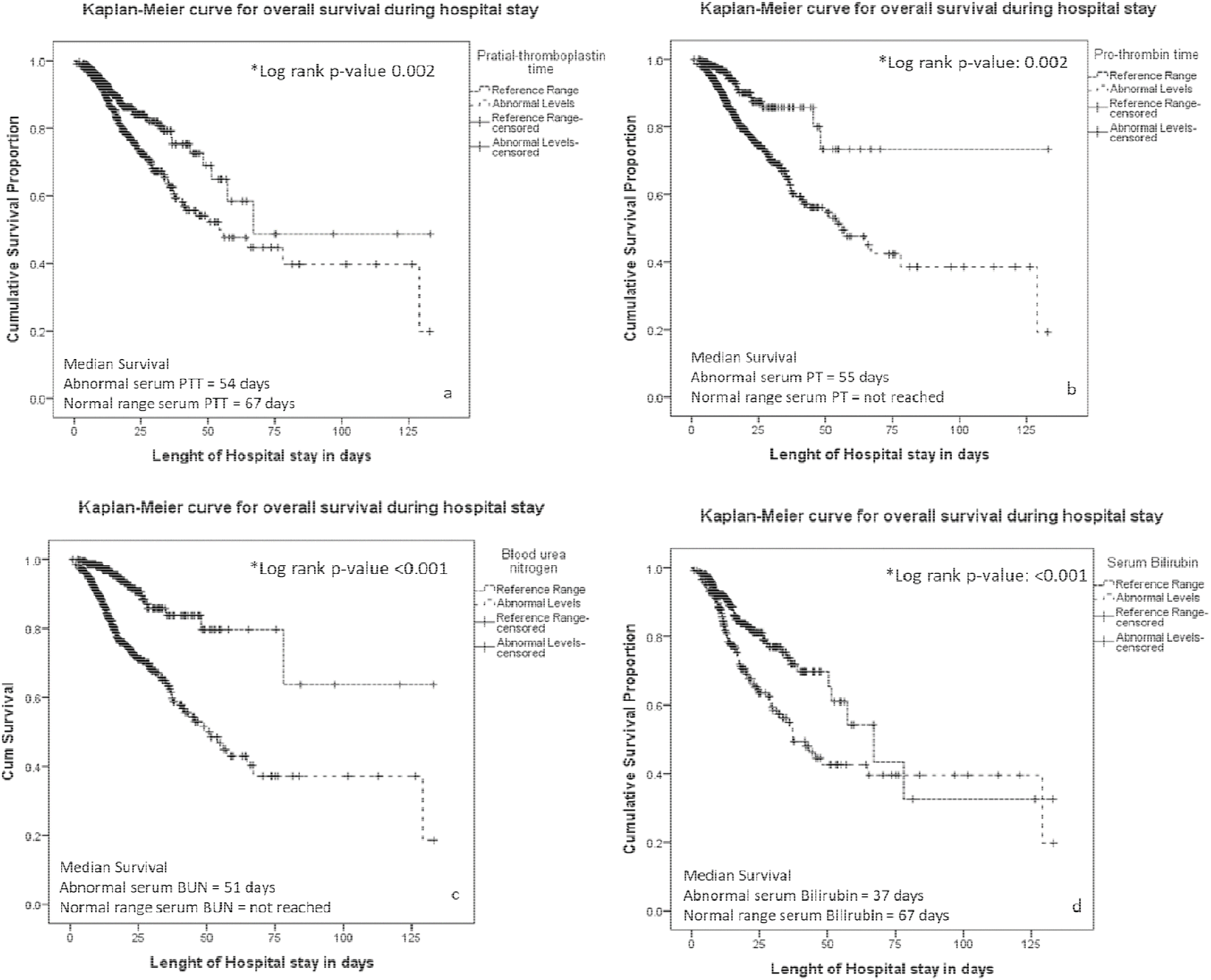
Kaplan-Meier Curves for a) abnormal serum PTT (MS 54 days vs 67 day), b) abnormal serum PT (MS 55 days vs DNRMS), c) abnormal serum BUN (MS 51 days vs MSDNR), d) abnormal serum Bilirubin (MS 37 days vs 37day). MS = median survival, DNRMS = did not reach median

**Supplemental Figure 4.**
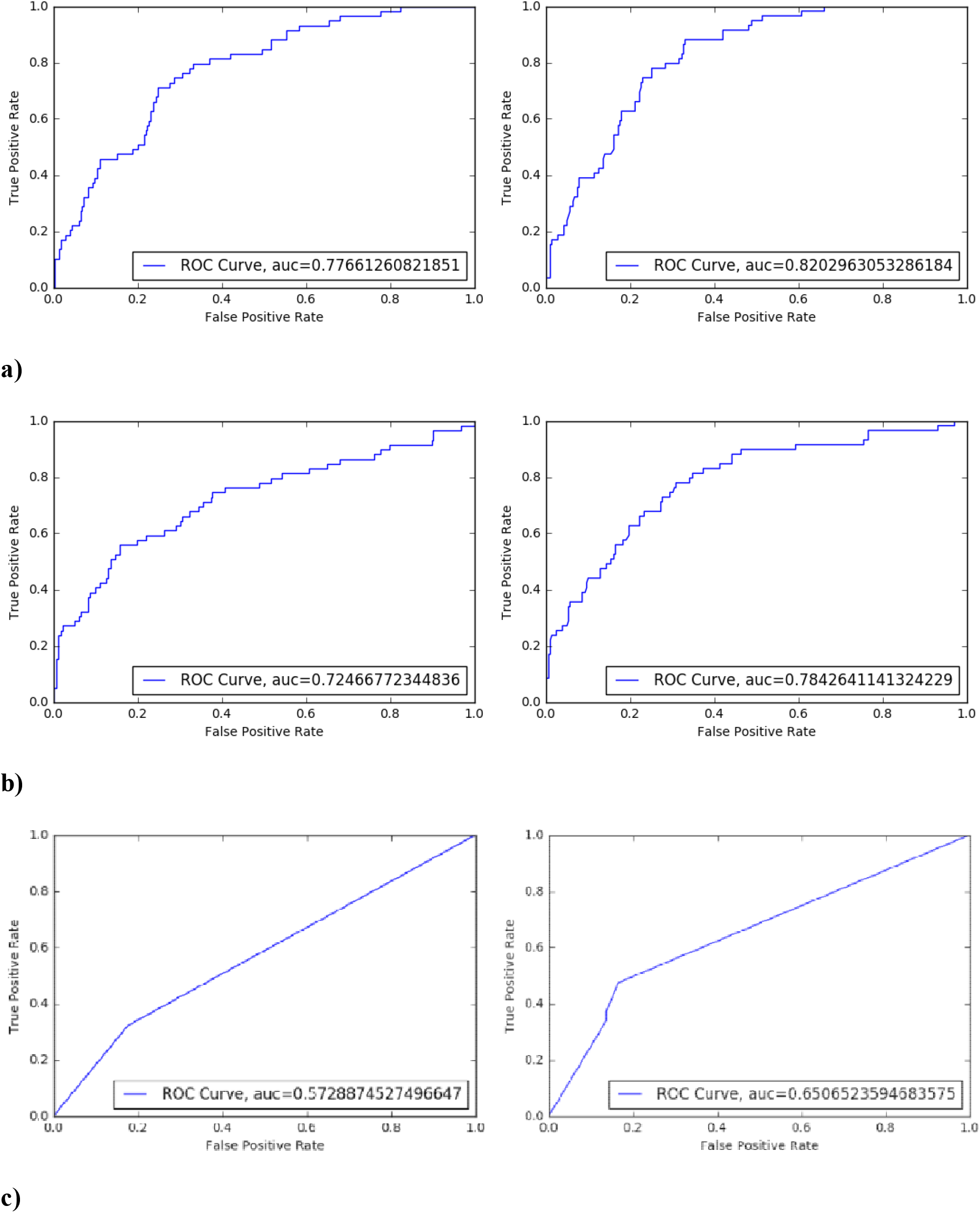
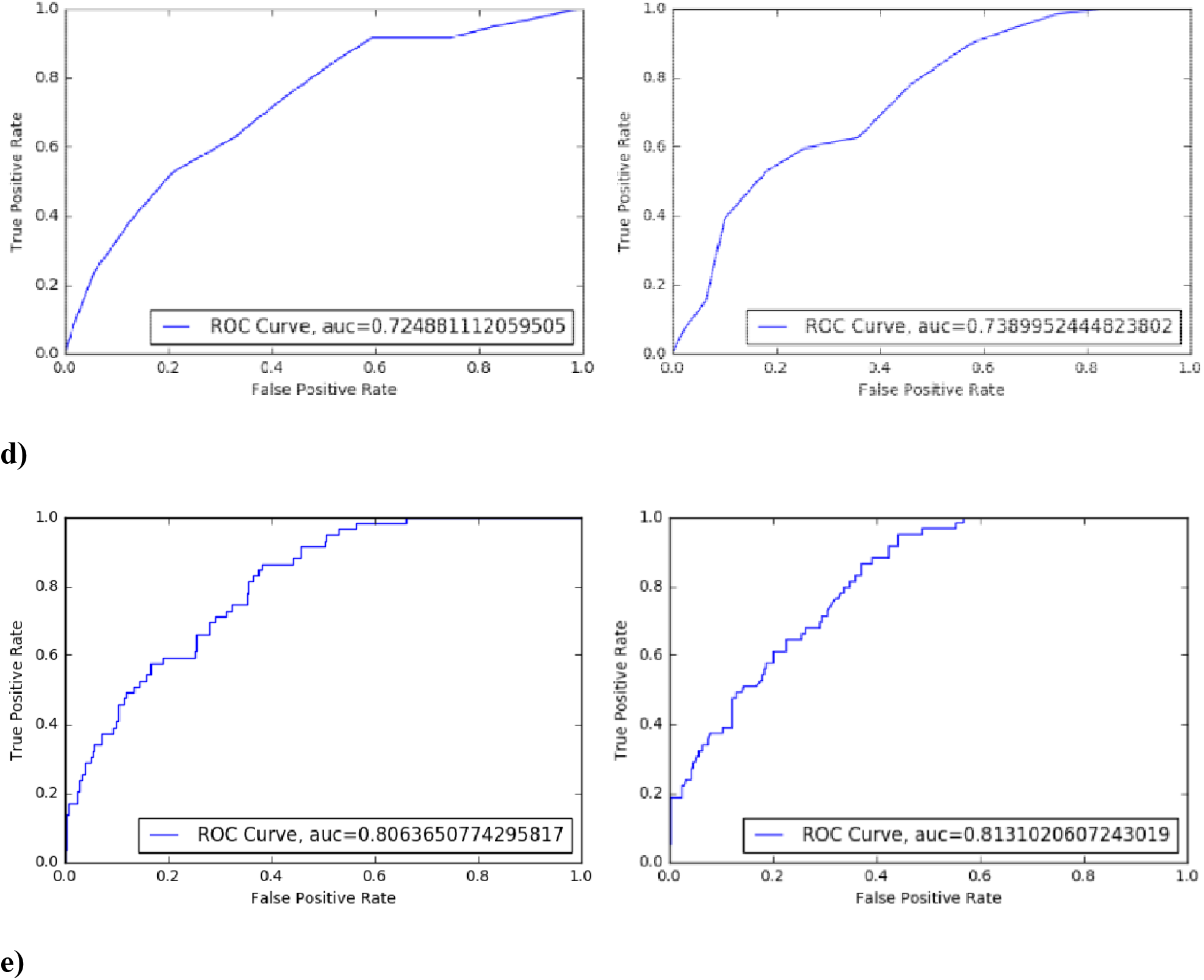
a) Left ROC: LDA Conventional Model (Baseline model), Right ROC: LDA Proposed. Improvement in AUC due to application of FLAIM architecture = 4.36%. b) Left ROC: GNB Conventional Model (Baseline model), Right ROC: GNB Proposed. Improvement in AUC due to application of FLAIM architecture = 5.96%. c) Left ROC: DT Conventional Model (Baseline model), Right ROC: DT Proposed. Improvement in AUC due to application of FLAIM architecture = 7.78%. d) Left ROC: KNN Conventional Model (Baseline model), Right ROC: KNN Proposed. Improvement in AUC due to application of FLAIM architecture = 1.41%. e) Left ROC: SVM (Lin) Conventional Model (Baseline model), Right ROC: SVM (Lin) Proposed. Improvement in AUC due to application of FLAIM architecture = 0.68%.

